# Impact of a Social Media Campaign on HIV-Related Stigma among Young Adults Living with HIV in Lima, Peru: A Sequential Explanatory Mixed Methods Study

**DOI:** 10.64898/2026.05.04.26352384

**Authors:** Samara Ruberg, Alyson Nunez, Milagros Wong, Marguerite Curtis, Yihan Shi, Hugo Sánchez, Eduardo Matos, Frine Samalvides, Kristin Kosyluk, Jerome T Galea, Renato Errea, Molly F. Franke

## Abstract

**Background:** Stigma remains a pervasive barrier to curbing the spread of human immunodeficiency virus (HIV) among adolescents and young adults in Lima, Peru. Social media offers a promising avenue for scalable, youth-centered stigma reduction, but few interventions have been rigorously evaluated in this context.

**Objective:** We evaluated the potential of a social media campaign to reduce perceived HIV-related stigma among young adults living with HIV. This involved a sequential explanatory mixed-methods study, including a randomized evaluation, followed by focus groups to understand the findings.

**Methods:** 150 young adults (aged 18-29 years) living with HIV (YLWH) were randomized to receive information on social media from one of the following: (1) the control account; (2) the control account and the social media campaign accounts (Instagram and TikTok); or (3) the control account, the campaign accounts, and the accounts of participating influencers. Perceived stigma was measured via pre- and post-campaign surveys using Spanish versions of the abridged Berger HIV Stigma Scale and the Stigma Stress Scale. Focus groups and interviews were conducted with a purposive sample of participants to contextualize quantitative results. Qualitative data were analyzed using Framework Analysis.

**Results:** Mean changes in HIV Stigma and Stigma Stress scores were small and not statistically significant. Post-hoc as-treated analyses supported these findings. Fidelity to intervention allocation was low to moderate, depending on the metric considered. Qualitative data suggested that the campaign positively impacted participants’ perceived stigma and that personal circumstances, crossover, frequency of exposure to content, and issues related to completing study questionnaires contributed to the lack of meaningful change in stigma scores.

**Conclusions:** While quantitative data did not support that exposure to a social media campaign led to meaningful reductions in HIV-related stigma, qualitative data suggested that the campaign had a positive impact and that limitations in the study design, together with external factors, may have obscured benefits in quantitative analyses.

## INTRODUCTION

In 2020, the Joint United Nations Programme on HIV/AIDS updated global targets for testing and successful treatment of human immunodeficiency virus (HIV) to be achieved by 2025.^1^ Also known as the “95-95-95” targets, 95% of all people living with HIV (PLWH) were to know their HIV status; 95% of all people with diagnosed HIV infection were to receive sustained antiretroviral therapy, and 95% of all people receiving antiretroviral therapy were to have viral suppression. These targets are not on track to be met globally;^2^ service delivery gaps remain in many regions of the world, and HIV-related stigma constitutes a major barrier to closing them. In Latin America, HIV-related stigma remains particularly prevalent and impacts national efforts to curb ongoing transmission and ensure access to HIV-related services.^3,4^ The harmful consequences of HIV-related stigma disproportionately impact adolescents and young adults,^5^ a population that is already at high risk of unfavorable HIV-related health outcomes.^6–8^

To improve the health of the most underserved communities, new and innovative strategies are required. Social media offers a mechanism for low-cost, far-reaching health communication. For example, applications such as Instagram and TikTok are internet-based platforms through which videos, memes, and other media are widely and inexpensively disseminated. These information-sharing platforms are particularly apt for populations with ready access to and experience using the internet and mobile devices, as is the case in Peru, where there were 24 million active social media user identities in a population of 34 million at the beginning of 2024.^9^ Education, through the contrasting of myths and facts, and contact, through the sharing of lived experience of a stigmatized identity, have proven to be effective approaches for reducing stigma, including internalized stigma,^10,11^ and recent investigations have demonstrated the success of social media-based interventions aiming to combat misinformation, mitigate stigma, and increase testing and treatment for diverse health conditions.^12–14^ Local influencer participation in public health social media campaigns has also proven to be an effective way of presenting accurate and culturally-aligned information about COVID-19 vaccination and transmission of bloodborne diseases.^15,16^ Lacking, however, are studies on how social media and influencers can be harnessed to impact HIV-related stigma.

We conducted a Spanish-language, community-engaged social marketing campaign that aimed to reduce HIV-related stigma among young people in Lima, Peru.^17^ The goal of the campaign was to address common myths and stereotypes regarding HIV, encourage empathy for PLWH, and normalize conversations around testing, prevention, and treatment.^18^ Local influencers participated by creating and sharing content to achieve these goals. Here, we report the results of research examining whether exposure to the campaign reduced perceived stigma among young adults living with HIV.

## METHODS

### Study Setting

There are an estimated 110,000 PLWH in Peru, with the highest prevalence in the nation’s capital, Lima.^19,20^ Peru has a concentrated HIV epidemic, with key populations of gay, bisexual, and other men who have sex with men (GBMSM) and transgender women (TGW) most affected.^19^ In 2024, adolescents and young adults (15-29) accounted for 47% of new diagnoses.^19^

Stigma is common among PLWH in Peru. In a 2023 survey, 76% of PLWH reported feelings of self-stigma, and 18% experienced discrimination from an HIV care provider.^4^ This stigma negatively impacts each step of the HIV care cascade in Peru, from testing to treatment to viral suppression, and hinders progress to global targets.^21–24^ In 2024, 87% of those living with HIV in Peru knew their status, 92% of those diagnosed were receiving treatment, and 83% of those receiving treatment had a suppressed viral load, presenting an opportunity to strengthen the HIV cascade through stigma reduction efforts.^20^

### The DiME Social Media Campaign

The DiME (in Spanish, “Tell Me”) social media campaign to address HIV-related stigma ran from July to December 2024 on Instagram and TikTok (@dime.peru).^17^ Both accounts were small at the start of the campaign, with no more than 600 followers each. A total of 49 videos were created by a content creation agency and the study team, informed by formative qualitative research with young PLWH (YPLWH), HIV advocates, and clinical providers.^18^ Additionally, seven highly recognizable paid local influencers (follower counts ranging from 117K to 23.8M) created 13 videos to post as part of the campaign, and 78 unpaid local influencers with more modest followings (ranging from 4.1K to 689K) were invited to share campaign content in their Instagram stories. Content sought to reduce HIV-related stigma through the combination of education and contact, combatting misinformation about HIV while promoting empathy and normalizing discussion of the condition.^18^ Video formats included scripted scenes, testimonials from YPLWH, and live interviews, among others.^17^ All videos were reviewed and approved by the study team and an advisory committee of stakeholders, including HIV advocates and YPLWH. Overall posting frequency was between two and three videos per week.

### Study Population

Participants were young adults living with HIV between the ages of 18 and 29 years who used social media (specifically, Instagram and/or TikTok) at least three times per week. They were recruited from two public sector health facilities where they received HIV care, Hospital Nacional Arzobispo Loayza and Hospital Nacional Cayetano Heredia, and from the NGO Epicentro, which offers HIV treatment and care services, with a focus on the LGBTQIA+ community.

### Study Design

We conducted a sequential explanatory mixed-methods study. First, we conducted a randomized controlled evaluation to evaluate the potential effectiveness of a community-engaged social media campaign to reduce HIV-related stigma among young adults. Results were then used to inform a subsequent qualitative study to facilitate interpretation of quantitative findings.

### Randomized Controlled Evaluation

Participants were randomized 1:1:1 to follow one or more social media accounts on Instagram and TikTok:(1) the control account (i.e. the Peruvian Ministry of Health [MINSA]) [Arm 1]; (2) the control account and the DiME campaign account [Arm 2]; (3) the control account, the DiME campaign account, and accounts of participating influencers [Arm 3]. Randomization, stratified by health facility, was performed using sequentially numbered, opaque, sealed envelopes. A randomization schedule was created using random permuted blocks of three different sizes. After randomization, participants were asked to follow their assigned social media accounts for the duration of the study. Participants in Arm 3 were asked to follow the control account, the DiME accounts, and seven top influencer accounts, and to select as many additional influencers from the provided list of 52 as they deemed appropriate, based on their interests.

### Data Collection

#### Quantitative

Participants completed a pre-randomization survey that included questions on sociodemographic characteristics, frequency and nature of social media usage, and perceived stigma and stigma stress (the primary outcomes). Participants were asked to self-report Instagram and TikTok usage and their handles. Stigma was assessed using Spanish versions of the abridged Berger HIV Stigma Scale and Stigma Stress scales. Both scales capture perceptions of stigma at the time of survey administration. The Socios En Salud Youth Advisory Board, comprised of 10 YPLWH, reviewed surveys for clarity and language.

The Berger HIV Stigma Scale uses a 4-point Likert scale to measure perceived stigma among individuals living with HIV and has been validated for use in Peru.^25,26^ The scale consists of four domains: Disclosure Concerns (ranging from 5 to 20 points), Negative Self-Image (ranging from 6 to 24 points), Concern with Public Attitudes (ranging from 5 to 20 points), and Enacted Stigma (ranging from 5 to 20 points). We excluded the Enacted Stigma subscale because it reflects the actions of others, who may not be exposed to the campaign content, toward PLWH; therefore, participants may be less likely to experience a change in this metric over the short term. The resulting 16-question HIV Stigma Scale score ranged from 16 to 64 points, with higher scores representing greater levels of perceived stigma.

The 8-question Stigma Stress Scale, used to measure stigma stress related to mental illness, was adapted for use in the context of HIV. In our Spanish translation, the phrase ‘mental illness’ was replaced with ‘HIV’ and ‘stigma’ was replaced with ‘stereotypes and prejudice’ for comprehension by participants. In keeping with the original version, a 7-point Likert scale was used to measure perceived stigma-related harm and coping resources.^27–29^ The mean coping resources score (items 5-8) was subtracted from the mean perceived harm score (items 1-4), yielding an overall composite stigma stress score ranging from -6 to 6, with positive scores indicating higher perceived stigma stress.

The pre-randomization survey was completed between 14 June and 8 July 2024. From October 2024 to January 2025, following the end of the intensive phase of the campaign, participants completed an abbreviated follow-up survey in which primary outcomes were reassessed. Participants self-reported whether they followed the assigned accounts, and the study team also used participant handles to track fidelity to the assigned study arm on a weekly basis.

#### Qualitative

A diverse group of participants from each study arm was purposively selected to participate in post-intervention focus groups or interviews. Focus group and interview guides were developed in response to preliminary analyses that yielded null quantitative results, and they explored exposure to campaign content, perceived stigma levels, potential external causes of stigma, and campaign impact (Interview Guide – Table S1 in Multimedia Appendix 1). Ms. M. Wong, a Peruvian nurse experienced in conducting qualitative research with YPLWH, conducted seven focus groups and 12 interviews with a total of 40 participants at the Socios En Salud office (focus groups) and virtually (interviews) from February to April 2025. Sessions lasted approximately 60-90 minutes and consecutive sessions were carried out until data saturation was reached. No relationship existed between Ms. Wong and participants prior to the sessions, and no other individuals were present during data collection. Participants were assigned to focus groups based on study arm, direction of stigma change (increase vs. decrease), and whether they followed the DiME account. Sessions were audio-recorded and transcribed. Notes were not taken during sessions, and transcripts were not returned to participants for review. No repeat interviews were conducted. We adhered to COREQ standards for qualitative reporting (Table S1 in Multimedia Appendix 2).

### Data Analysis

#### Quantitative

Descriptive statistics, including means, standard deviations (SD), medians, and quartiles, were used to describe the data. Primary analysis was intention-to-treat. Changes in perceived HIV stigma and stigma stress were calculated and examined for normality. Difference-in-differences relative to the control were assessed using two-sample paired t-tests, and we conducted a secondary analysis using ANCOVA, adjusting for baseline imbalances in age, sex assigned at birth, sexual orientation, gender identity, education level, and months since diagnosis. Secondary as-treated analyses re-classified participants into two arms based on whether they followed the DiME account on either platform. For the first analysis, we classified campaign exposure based on data collected by the study team throughout the campaign, identifying which participant handles ever followed the DiME account on either platform, and compared changes using ANCOVA. We repeated this analysis using self-reported follow data. As-treated analyses were both unadjusted and adjusted for baseline covariate differences in age, sex assigned at birth, sexual orientation, gender identity, education level, and months since diagnosis. In a post hoc analysis, we pooled data across study arms to assess overall changes in stigma during the campaign. Data from pre- and post-intervention surveys were analyzed using RStudio and Statistical Analysis Software (SAS) version 9.4.

#### Qualitative

Focus group and interview transcripts were deidentified and uploaded into Dedoose for qualitative analysis. Pseudonyms were assigned to distinguish participants. Framework Analysis methodology guided data analysis. A preliminary codebook based on the semi-structured interview guide was created, and *de novo* codes were added as needed (Codebook – Table S1 in Multimedia Appendix 1). Two study team members coded each of the 19 transcripts independently. A third member reviewed the applied codes to resolve discrepancies and reach consensus. Data were then grouped into emergent themes for analysis. Participants did not provide feedback on study findings.

#### Sample size calculations for randomized evaluation

The study was powered to detect a change in the HIV Stigma Scale. Assuming 10% attrition (i.e., a final analytic cohort of 45 participants per arm), a sample size of 135 would provide 80% power to detect a difference-in-difference of +/-2.4 (type I error: 0.05, standard deviation:4).

### Ethical Considerations

The Vía Libre Bioethics Committee in Peru, the Ethics Committees at participating hospitals, and the Longwood Campus Institutional Review Board of Harvard Medical School approved this research. The randomized evaluation was classified as exempt under the Common Rule Category 3, which covers benign behavioral intervention in adults.^30^ Participants in focus groups and interviews provided written informed consent. All participants received reimbursement for transportation costs and 50 soles (15 USD) for each study encounter (i.e., surveys, focus group or interview).

## RESULTS

### Participant Characteristics

Of the 150 participants enrolled and randomized, 48 from Arm 1 (98%), 47 from Arm 2 (96%), and 46 from Arm 3 (88%) completed both the pre- and post-intervention surveys and were included in the analysis (Figure 1). Respondents ranged in age from 18 to 29 years (mean = 25.7; Table 1). Most were assigned male at birth (77%), identified as men (70%), and were born in Peru (87%). Half of all participants reported homosexual sexual orientation (54%) and listed high school as the highest level of education attained (53%). Most participants reported that they were taking antiretroviral therapy at enrollment (97%) and had previously disclosed their HIV diagnosis to at least one other individual (90%). Median time since HIV diagnosis was 68 months (Q1 = 35, Q3= 89, range = 0.5-288). Regarding social media usage, 58% reported using Instagram and 76% reported using TikTok seven days per week. Participants spent a median of two hours per day on Instagram and three hours per day on TikTok. A chance imbalance in the distribution of sex assigned at birth (ranging from 12.8% in arm 2 to 32.6% in arm 3) resulted in other imbalances in correlated variables (Table 1).

**Table 1.** Baseline characteristics of study participants.

| Characteristic | Overall<br>n (%)* | Control<br>Arm 1 (N = 48)<br>n (%)* | DiME<br>Arm 2 (N = 47)<br>n (%)* | DiME and<br>Influencers<br>Arm 3 (N = 46)<br>(%)* |
| --- | --- | --- | --- | --- |
| **Age, mean (SD)** | 25.7 (2.9) | 25.4 (2.9) | 26.2 (2.6) | 25.6 (3.0) |
| **Sex assigned at birth** |  |  |  |  |
| Female | 31 (22.0) | 10 (20.8) | 6 (12.8) | 15 (32.6) |
| Male | 109 (77.3) | 38 (79.2) | 41 (87.2) | 30 (65.2) |
| Intersex | 0 (0.0) | 0 (0.0) | 0 (0.0) | 0 (0.0) |
| Other | 1 (0.71) | 0 (0.0) | 0 (0.0) | 1 (2.2) |
| **Peruvian** | 123 (87.2) | 45 (93.8) | 38 (80.9) | 40 (87.0) |
| **Highest level of education** |  |  |  |  |
| Primary | 5 (3.6) | 1 (2.1) | 0 (0.0) | 4 (8.7) |
| High school | 75 (53.2) | 29 (60.4) | 24 (51.1) | 22 (47.8) |
| Trade school | 31 (22.0) | 9 (18.8) | 11 (23.4) | 11 (23.9) |
| University | 29 (20.6) | 9 (18.8) | 11 (23.4) | 9 (19.6) |
| Master's degree or higher | 1 (0.7) | 0 (0.0) | 1 (2.1) | 0 (0.0) |
| **Sexual orientation** |  |  |  |  |
| Heterosexual | 46 (32.6) | 16 (33.3) | 12 (25.5) | 18 (39.1) |
| Homosexual | 76 (53.9) | 26 (54.2) | 26 (55.3) | 24 (52.2) |
| Bisexual | 17 (12.1) | 4 (8.3) | 9 (19.2) | 4 (8.7) |
| Pansexual | 1 (0.7) | 1 (2.1) | 0 (0.0) | 0 (0.0) |
| Other | 1 (0.7) | 1 (2.1) | 0 (0.0) | 0 (0.0) |
| **Gender identity** |  |  |  |  |
| Woman | 32 (22.7) | 11 (22.9) | 7 (14.9) | 14 (30.4) |
| Man | 99 (70.2) | 35 (72.9) | 39 (83.0) | 25 (54.4) |
| Transgender woman | 4 (2.8) | 1 (2.1) | 0 (0.0) | 3 (6.5) |
| Transgender man | 1 (0.7) | 0 (0.0) | 0 (0.0) | 1 (2.2) |
| Gender fluid | 1 (0.7) | 1 (2.1) | 0 (0.0) | 0 (0.0) |
| Non-binary | 3 (2.1) | 0 (0.0) | 1 (2.1) | 2 (4.4) |
| Other | 1 (0.7) | 0 (0.0) | 0 (0.0) | 1 (2.2) |
| **Months since HIV diagnosis, median (Q1, Q3, min, max) (N = 140)** | 68.2 (34.6, 88.7, 0.5, 288.1) | 67.5 (26, 87.7, 0.6, 282.8) | 73.6 (41.7, 89.6, 0.5, 261.5) | 70.8 (32.7, 96, 0.6, 288.1) |
| **Taking antiretroviral treatment** | 136 (96.5) | 46 (95.8) | 46 (97.9) | 44 (95.6) |
| **Disclosed HIV diagnosis to ≥ 1 person** | 127 (90.1) | 47 (97.9) | 43 (91.5) | 37 (80.4) |
| **Experienced stigma or discrimination within the last 12 months** | 29 (20.6) | 9 (18.8) | 10 (21.3) | 10 (21.7) |
| **Self-description of health** |  |  |  |  |
| Excellent | 25 (17.7) | 6 (12.5) | 13 (27.7) | 6 (13.0) |
| Very good | 39 (27.7) | 12 (25.0) | 13 (27.7) | 14 (30.4) |
| Good | 50 (35.5) | 18 (37.5) | 16 (34.0) | 33 (23.4) |
| Regular | 26 (18.4) | 11 (22.9) | 5 (10.6) | 10 (21.7) |
| Bad | 1 (0.7) | 1 (2.1) | 0 (0.0) | 0 (0.0) |
| **Has a smartphone** | 131 (92.9) | 46 (95.8) | 40 (85.1) | 45 (97.8) |
| **Uses TikTok daily**<br>(N = 134) | 102 (76.1) | 31 (70.5) | 35 (76.1) | 36 (81.8) |
| **Hours spent per day on TikTok, median (Q1, Q3)**<br>(N = 140) | 3 (2, 5.25) | 4 (2, 5) | 3 (2, 6) | 3 (2, 5) |
| **Uses Instagram daily**<br>(N = 138) | 80 (58.0) | 23 (50.0) | 29 (63.0) | 28 (60.9) |
| **Hours spent per day on Instagram, median (Q1, Q3)**<br>(N = 138) | 2 (1, 3) | 3 (2, 3.5) | 2 (1, 3) | 2 (1, 3) |

\*Unless otherwise noted.

**Figure 1.**
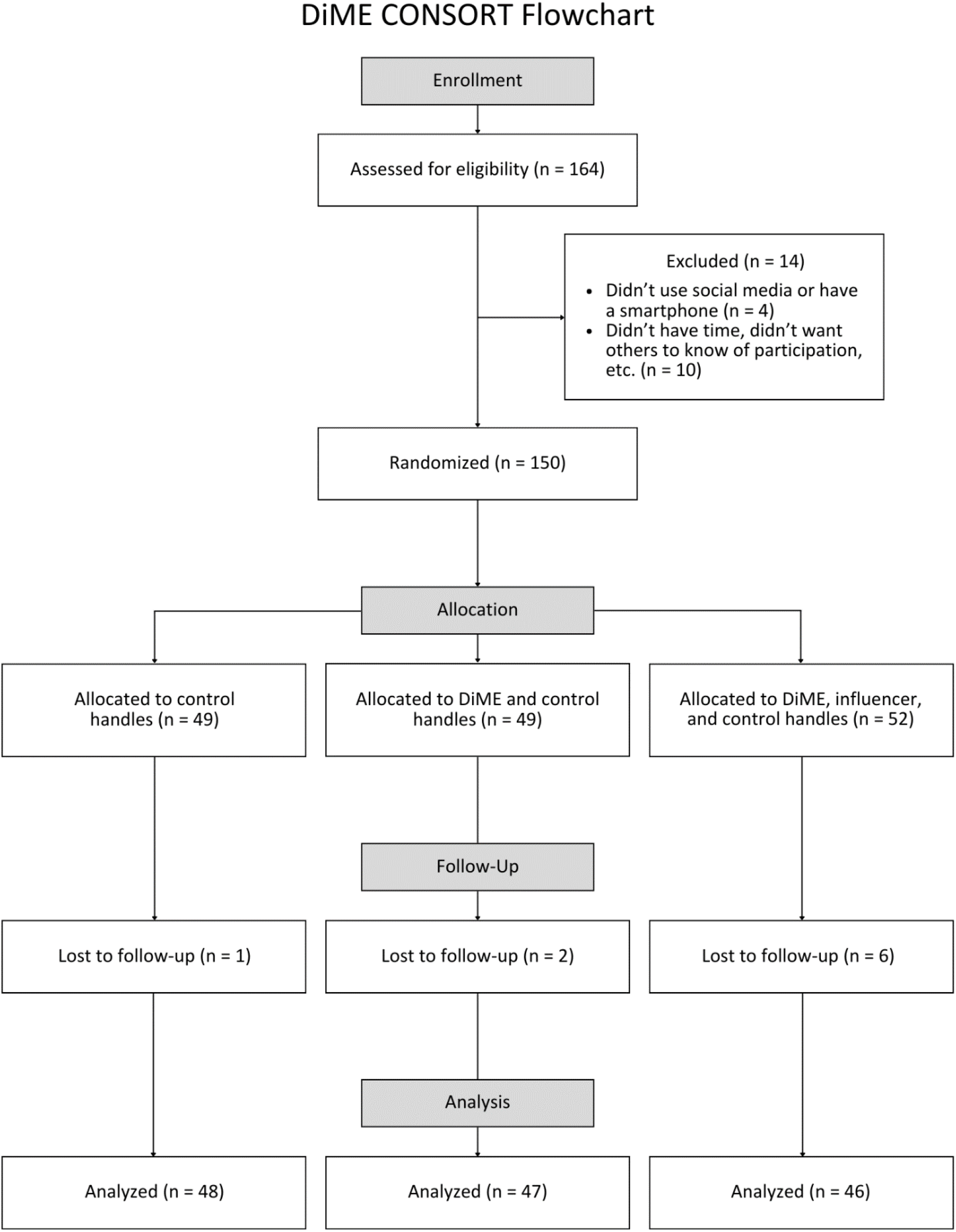
CONSORT diagram.

### Baseline HIV Stigma and Stigma Stress Scores

Mean overall HIV Stigma scores at baseline ranged from 40.0 to 42.1 (Table 2). The Disclosure Concerns and Concern with Public Attitudes subscales, both with possible values ranging from 5 to 20, had the highest mean scores, ranging from 14.4 to 15.9 and 14.0 to 15.1, respectively. Mean scores for the Negative Self-Image subscale, with possible values of 6 to 24, ranged from 11.0 to 11.2. Mean Stigma Stress scores at baseline ranged from -2.1 to -2.9 (range of possible values -6 to 6). Cronbach’s alpha was 0.86 for the HIV Stigma Scale, 0.89 for Perceived Harm Stigma Stress Subscale, and 0.82 for the Perceived Coping Resources Stigma Stress Subscale.

**Table 2.**
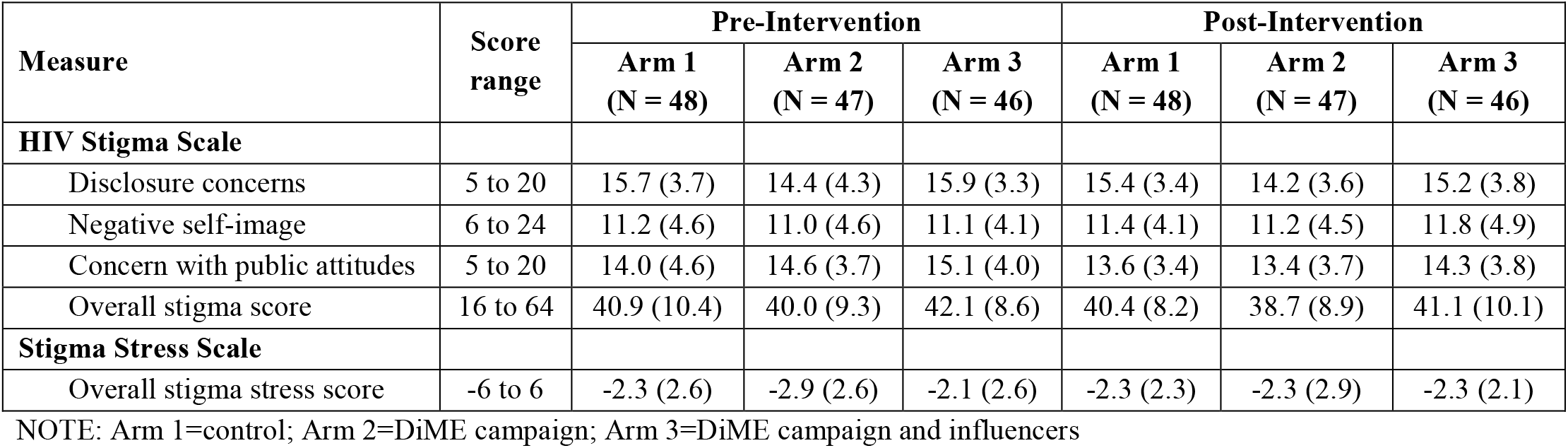
Mean HIV Stigma and Stigma Stress scores.

| Measure | Score range | Pre-Intervention |  |  | Post-Intervention |  |  |
| --- | --- | --- | --- | --- | --- | --- | --- |
|  |  | Arm 1<br>(N = 48) | Arm 2<br>(N = 47) | Arm 3<br>(N = 46) | Arm 1<br>(N = 48) | Arm 2<br>(N = 47) | Arm 3<br>(N = 46) |
| **HIV Stigma Scale** |  |  |  |  |  |  |  |
| Disclosure concerns | 5 to 20 | 15.7 (3.7) | 14.4 (4.3) | 15.9 (3.3) | 15.4 (3.4) | 14.2 (3.6) | 15.2 (3.8) |
| Negative self-image | 6 to 24 | 11.2 (4.6) | 11.0 (4.6) | 11.1 (4.1) | 11.4 (4.1) | 11.2 (4.5) | 11.8 (4.9) |
| Concern with public attitudes | 5 to 20 | 14.0 (4.6) | 14.6 (3.7) | 15.1 (4.0) | 13.6 (3.4) | 13.4 (3.7) | 14.3 (3.8) |
| Overall stigma score | 16 to 64 | 40.9 (10.4) | 40.0 (9.3) | 42.1 (8.6) | 40.4 (8.2) | 38.7 (8.9) | 41.1 (10.1) |
| **Stigma Stress Scale** |  |  |  |  |  |  |  |
| Overall stigma stress score | -6 to 6 | -2.3 (2.6) | -2.9 (2.6) | -2.1 (2.6) | -2.3 (2.3) | -2.3 (2.9) | -2.3 (2.1) |

NOTE: Arm 1=control; Arm 2=DiME campaign; Arm 3=DiME campaign and influencers

### Changes in HIV Stigma and Stigma Stress and Differences by Study Arm

Following the intervention period, the mean change in HIV Stigma scores was -0.46 in Arm 1, -1.36 in Arm 2, and -1.00 in Arm 3 (difference-in-difference p-value, with Arm 1 as reference: 0.60 and 0.77, respectively) (Table 3). Mean change scores for the Disclosure Concerns subscale were -0.29 in Arm 1, - 0.29 in Arm 2 (p-value=0.96), and -0.74 in Arm 3 (p-value=0.58); for the Concern with Public Attitudes subscale were -0.38 in Arm 1, -1.23 in Arm 2 (p-value=0.29), and -0.87 in Arm 3 (p-value=0.56). Negative Self-Image mean change scores were 0.21 in Arm 1, 0.13 in Arm 2 (p-value=0.93), and 0.61 in Arm 3 (p-value=0.68). On the Stigma Stress Scale, the mean change following the intervention was 0.05 in Arm 1, 0.56 in Arm 2 (p-value=0.40), and -0.2 in Arm 3 (p-value=0.64) (Figure 2). There were no substantive differences after adjusting for baseline covariate imbalances (Table S1 in Multimedia Appendix 3).

**Table 3.** Mean change in HIV Stigma and Stigma Stress scores.

| Measure | Arm 1 (N=48) | Arm 2 (N=47) |  | Arm 3 (N=46) |  |
| --- | --- | --- | --- | --- | --- |
|  | mean (95% CI) | mean (95% CI) | p-value* | mean (95% CI) | p-value* |
| **HIV Stigma Scale** |  |  |  |  |  |
| Disclosure concerns | -0.29 (-1.52, 0.94) | -0.26 (-1.28, 0.77) | 0.96 | -0.74 (-1.79, 0.31) | 0.58 |
| Negative self-image | 0.21 (-1.20, 1.62) | 0.13 (-1.00, 1.26) | 0.93 | 0.61 (-0.78, 1.99) | 0.68 |
| Concern with public attitudes | -0.38 (-1.62, 0.88) | -1.23 (-2.28, -0.18) | 0.29 | -0.87 (-2.00, 0.26) | 0.56 |
| Overall stigma score | -0.46 (-3.21, 2.29) | -1.36 (-3.41, 0.69) | 0.60 | -1.00 (-3.56, 1.56) | 0.77 |
| **Stigma Stress Scale** |  |  |  |  |  |
| Overall stigma stress score | 0.05 (-0.57, 0.66) | 0.56 (-0.50, 1.62) | 0.40 | -0.20 (-1.06, 0.67) | 0.64 |

NOTE: Arm 1=control; Arm 2=DiME campaign; Arm 3=DiME campaign and influencers
\*p-value testing difference in differences, with Arm 1 as the reference group

**Figure 2.**
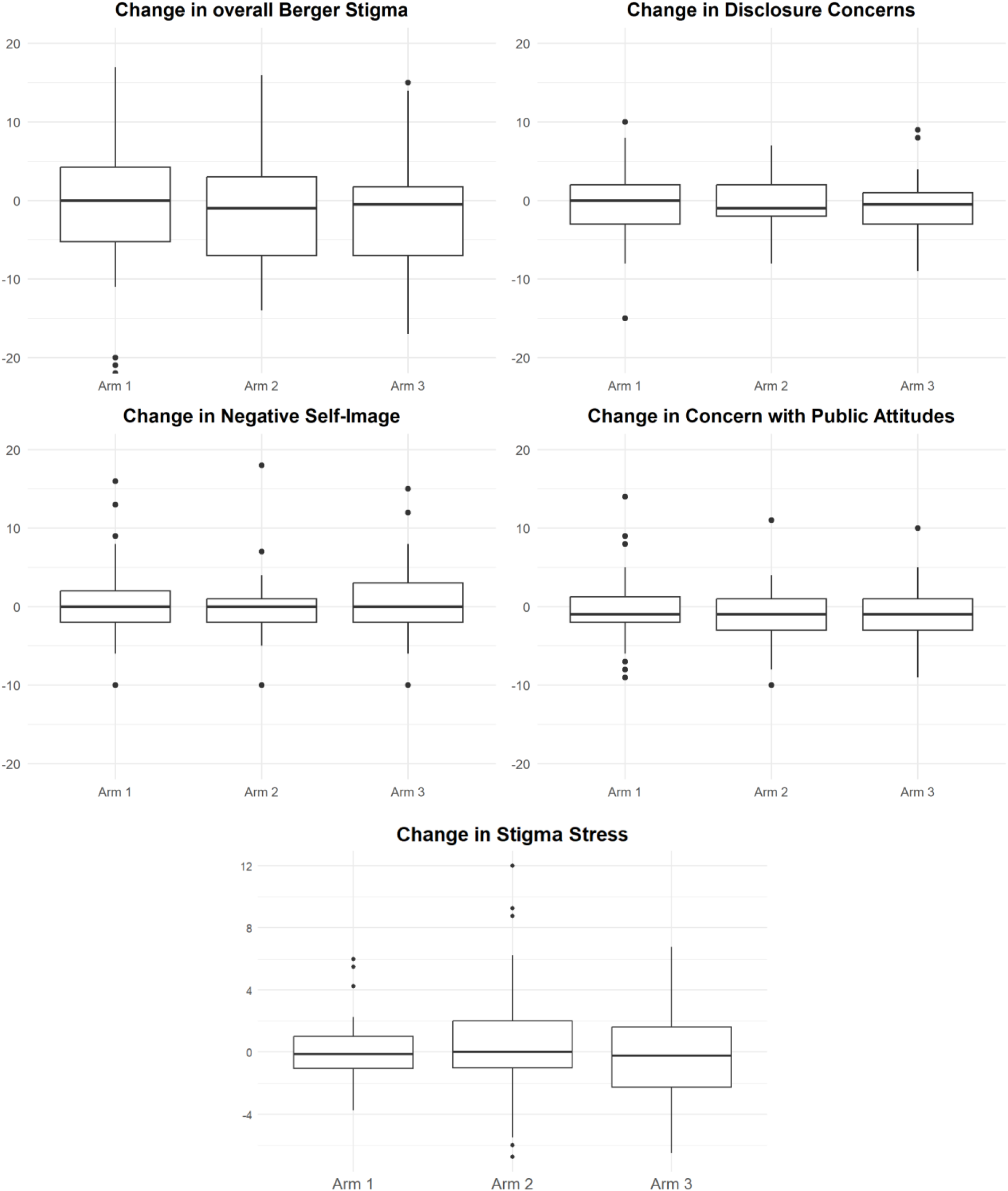
Boxplots showing mean change in overall HIV Stigma Scale, subscales, and Stigma Stress Scale by arm. The central line indicates mean change. Boxes span the 25^th^ to 75^th^ percentile, and whiskers show the minimum/maximum values, excluding outliers (dots). Two outliers (Arm 1=33, Arm 3=26) for change in overall HIV stigma not shown.

### Fidelity to Study Arm

According to the study team’s assessment of handle following data, 41 (29%) participants followed the DiME account on either platform (Arm 1: 3 (6%), Arm 2: 30 (64%), Arm 3: 8 (17%)). According to self-reported data, 110 (78%) participants followed the account (Arm 1: 26 (54%), Arm 2: 43 (91%), Arm 3: 41 (89%)). The distribution of characteristics across study arms in as-treated analyses are shown in Tables S2 and S3 in Multimedia Appendix 3.

### Secondary and Post Hoc Analyses

In the first as-treated analysis, with exposure classified based on the study team’s tracking of handles, mean change in overall HIV Stigma was -0.47 (95% CI: -2.12, 1.18) for the control arm and -2.07 (95% CI: - 4.76, 0.61) for those who followed the DiME account on either platform (Table S4 in Multimedia Appendix 3). Mean change in the Disclosure Concerns subscale was -0.27 (95% CI: -1.02, 0.48) and -0.80 (95% CI: -1.98, 0.37); in the Concern with Public Attitudes subscale was -0.67 (95% CI: -1.44, 0.10) and -1.20 (95% CI: -2.46, 0.07); and for the Negative Self-Image subscale was 0.47 (95% CI: -0.45, 1.39) and -0.07 (95% CI: -1.33, 1.18), for control and intervention arms, respectively. The mean change in Stigma Stress score was 0.27 (95% CI: -0.32, 0.85) in the control arm and -0.16 (95% CI: -1.09, 0.76) in those who followed the account. Results were similar when adjusting for baseline differences (Table S5 in Multimedia Appendix 3). The second as-treated analysis, with exposure determined by participant self-report, yielded results similar to the first, with no meaningful differences observed in either the unadjusted or adjusted analyses (Tables S6 and S7 in Multimedia Appendix 3).

In post hoc analyses pooling across all arms, the Concern with Public Attitudes subscale showed a statistically significant reduction of -0.82 (95% CI: -1.47, -0.17; p = 0.014) between pre- and post-interviews (Table S8 in Multimedia Appendix 3). No other statistically significant reductions were observed.

### Qualitative Analysis

Qualitative results provided important context for interpreting the quantitative findings. Participants exposed to the campaign reported that its content positively impacted their perceived stigma. They also noted that personal circumstances, frequency of exposure to content, and issues with completing study questionnaires may have influenced the observed lack of meaningful change in the stigma score.

#### A positive impact on stigma

Most study participants who viewed campaign content perceived the DiME campaign as having a positive impact on their perceived stigma. Following exposure to campaign content, participants described increased empowerment and HIV literacy. Several mentioned that the informal and personable nature of the videos conveyed a sense of calm and comfort, contrasting with the HIV-related communications to which they were accustomed, which tended to focus on testing and treatment. Participants identified with the situations and testimonials presented in the content and expressed resonance with their own journeys of self-acceptance of their diagnoses. They appreciated that videos reinforced familiar concepts while also introducing new information, which in some cases led to additional self-directed research. One participant explained that after viewing the videos, he was able to speak openly with his partner about his diagnosis. Others shared content with friends and family, thereby relieving themselves of the burden of personally explaining the information.

> *You treat the topic so light-heartedly… it makes you feel at peace, it gives you confidence. When people give their testimonies, I feel like “oh I could be there too*.*” So you really identify with what you’re seeing, and they’re stories that are sometimes very similar to yours and I really like that. (Bryan, Arm 1)*
>
> *It’s still hard for me sometimes, because I’m the kind of person who has trouble telling my family about my diagnosis. So yes, [living with HIV] does still affect me a bit. But the videos I’ve seen have helped me a lot, at least to tell my partner. Before I was scared, when I had to go to the hospital and tell the doctors, because I thought they would discriminate against me. But not now, now I just tell them. (Luciano, Arm 2)*

The campaign’s most viral video (388K views), depicting a young gay couple in a serodiscordant relationship, was described as especially impactful. Participants recalled feelings of despair upon learning their diagnosis, thinking they would never fall in love or have a relationship. They reported that this video, in particular, helped them and others understand that the condition is not an impediment to a healthy relationship.

> *The video I most identified with was the one with a serodiscordant couple because I’ve been in a relationship like that… it helps people. For example, at one point I felt like “I’m never going to fall in love, my life is over, I’m not going to be happy, I’m never going to have a relationship again*.*” So I think [the video] helps a lot. (Juan, Arm 2)*

#### Intensity of exposure to the campaign varied among those randomized to intervention arms

Participant exposure to campaign content varied: some participants randomized to intervention arms continued to follow the DiME account throughout the study period, whereas others reported having stopped after changing their social media account names or cellphone numbers. The majority saw DiME content only when it appeared in their feeds; a few reported intentionally opening a DiME account to view content, sometimes influenced by their commitment to the study. Additional factors contributing to limited exposure included inconsistent access to reliable internet and time constraints due to work and family responsibilities.

> *I waited for [the videos] to appear. I didn’t say, “let’s go look at DiME to see what it is*.*” No, no, it wasn’t like that. (Daniel, Arm 3)*
>
> *At the beginning, at least the first week, I was very attentive to posts. But later, because of work, I’m traveling constantly… and I don’t have [cellphone] signal [for weeks]. It’s a little frustrating. (Mario, Arm 2)*

#### Infidelity to assigned study arms was observed

Several control participants confirmed they had followed the campaign account. For example, one participant explained that he had searched for the account after seeing the DiME name in study materials left behind by another participant. Conversely, not all participants assigned to Arms 2 or 3 followed the DiME account, and some participants in Arm 3 reported following only some designated influencers or none at all. Most mentioned that the decision not to follow was due to busy personal schedules, though one participant explained that because he frequently saw DiME videos in his feed, he thought he was following the account when he in fact was not. Another described the decision to follow someone online as highly personal and grounded in real connections.

> *I didn’t sit down to watch all the videos. With everything that I have going on with my kids here, I didn’t follow the account. (Elisa, Arm 2)*
>
> *To follow someone, I have to know them personally. I have to have some connection. (Agustín, Arm 3)*

#### External factors impacted perceptions of stigma during the campaign period

Participants identified several external factors that may have impacted levels of perceived stigma and stigma stress. Some described specific instances of discrimination by family members, friends, coworkers, and healthcare professionals, and recognized that they continued to struggle with self-stigma (the internalization of misinformation and negative public attitudes) and the fear of what others might think. In contrast, others experienced support and acceptance from those to whom they disclosed their diagnosis during the study period, with one peer even becoming a participant’s emergency contact at the hospital. Several participants also highlighted how mood affects their interpretations of questions, explaining, for example, that a bad mood could influence responses. Finally, several participants suggested that simply being invited to participate in the study and knowing there was a social media account dedicated to reducing HIV-related stigma contributed to reductions in their own perceptions of stigma.

> *I mean I can say “today I’m ok, but not tomorrow,” but that’s me, it’s like an internal battle with myself every day. (Juan, Arm 2)*
>
> *Since being invited to participate, I’m more aware of the kind of information I’ve been receiving, and if at some point some person or group rejects me, it’s because of their lack of knowledge. It’s not my fault. So, I feel better knowing that, and that’s what’s started to come out for me. Because there are, and continue to be, platforms dedicated to providing more information about our experiences. (Francisco, Arm 2)*

#### Stigma scales may have not have adequately captured experiences of stigma

Participants noted that factors associated with survey completion may also have influenced responses. Some were unsure how to respond or what to think when completing the questions because of language perceived to be overly scientific. Others reported completing the survey quickly, needing to resume daily activities.

> *But let’s be honest, when you take a survey, sometimes it’s not because you didn’t understand but because there are so many similar answers. Or sometimes it’s the time, you want to leave quickly so you mark similar answers. (Jhonatan, Arm 2)*

Additional illustrative quotes are found in Table 4.

**Table 4.** Exemplary quotes: campaign impact, exposure to campaign, external factors, stigma questionnaire.

|  |  |
| --- | --- |
| The DiME campaign positively impacted stigma | *I felt like I was becoming more informed. I started to see that there were myths, that there are different prevention methods, etc. so more than anything it was a learning experience for me. (Jesús, Arm 3)*<br>*Even though they were things I already knew, [the videos] help me remember, they help me see things more dynamically and they help me want to learn more in some ways. That’s what this type of content encourages. (Benjamín, Arm 2)*<br>*The videos also helped me a lot. One video I saw, I said, “Oh ok, he has this condition,” in the video where he says ‘I have [HIV], my life is normal. I can work, it doesn’t mean that I can’t work’. So I said, “if he can do all this [with HIV] why can’t I? I also have to be able to. I have to do it. I have to get it out of my head that I can’t. I have to push that aside.” So, little by little. (Maritza, Arm 1)*<br>*I know someone who’s in a serodiscordant relationship, and I thought about that person, and I said, “Wow, how cool! If he saw this [video], he would feel really good, I think.” So yes, it’s a comfort to me in some ways. (Benjamín, Arm 2)* |
|  | *The video about the couple is going to help a lot because there are people not living with HIV who think that if they're with someone with HIV, they're going to get it. So I think those videos help a lot, influencing people to see that they can be in a relationship with someone living with HIV, that with the proper care and condoms there's no risk. (Luciano, Arm 2)* |
| Intensity of exposure to campaign varied | *I don't regularly see the videos in my feed. But when I remember, "oh what have they posted?" I go check. But it's also because of the commitment I made to the study. (Juan, Arm 2)* |
| Crossover between study arms was observed | *I think I started following because in the brochure that they gave me I think the [DiME] name was there. (Josué, Arm 1)*<br>*What happens is that sometimes the page comes up, or you search for it, you're watching the video, and since you're watching you're consuming content on the page. You give it a like but forget to follow. Because you're liking it, the app shows you more and more videos, so you think you're following. That's what happened to me. I didn't follow, but I was watching. (Andrés, Arm 2)* |
| External factors impacted perceptions of stigma | *I do think [my stigma levels] went down, but it was because I had been judged for my diagnosis in a relationship I was in and was feeling down. That was when I completed the first survey; the relationship then ended. I met someone else, and that person helped me see that it wasn't me, it wasn't my decision. And they didn't make me feel bad. (Leo, Arm 2)*<br>*At that time I had some emotional problems that were getting me down, not because of my diagnosis but for other reasons. But then at the second meeting [for follow-up], I felt more sure of myself, I didn't feel disoriented or down, I was stable so to speak, not like at the beginning. Everyone always has their highs and lows. (Josué, Arm 1)*<br>*One survey isn't going to be the same as another, how you show up that day to take the survey has a lot to do with it, or how you read it, or how you perceive it. In the first interview I might say I'm doing really well, I haven't experienced any stigma, everything's been fine, because during those days I'd been ok. But on the second survey, in those days, if I've experienced some discrimination or someone said something, or I heard a comment from my family or friends, this can influence the response when completing the questionnaire.... (Cristóbal, Arm 2)* |
| Experience completing stigma questionnaires may have influenced responses | *For example, the survey questions and the terms used may be drawn from scientific studies. And we're used to colloquial speech. We can't interpret it very well. That's worked against us sometimes. (Andrés, Arm 2)* |

## DISCUSSION

This study sought to assess whether exposure to a social media campaign, which included influencer-generated content, reduced HIV-related stigma among YPLWH in Lima, Peru. Quantitative data did not support an impact of the campaign on HIV-related stigma, though a number of limitations to the study design may have obscured differences across study arms. Conversely, qualitative data supported the campaign’s potential to contribute positively to stigma reduction and self-esteem among viewers. Several YPLWH described how seeing their peers leading active lives in DiME videos helped shift their mindset to one of “if they can do it while living with HIV, why can’t I?” In this way, some content may have acted as a form of unidirectional social support. Others reported learning new information regarding HIV-related myths and stereotypes. The upbeat, personable nature of DiME content was highlighted as a welcome departure from typical HIV-focused social media content, which was perceived to be primarily centered on testing and treatment.

Evaluating the efficacy of social media campaigns is inherently challenging. Observational designs are susceptible to bias arising from underlying time trends and from differences between users who consume particular content and those who do not. Randomized controlled evaluations, such as the one presented here, are often viewed as more rigorous; however, in the context of a social media intervention, maintaining control is challenging given social media’s widespread reach. This was evident in low rates of fidelity to the assigned interventions, which were difficult to assess and highly variable across assessment methods (self-report versus follow-up data). Some controls sought out content, whereas others may have encountered content serendipitously. This may have occurred as the account rapidly gained followers during the campaign and generated several viral posts (the top post was viewed more than 388K times). It may also have occurred due to the ways platforms promote “suggested content” based on existing networks and engagement. At the same time, a substantial number of participants in arms 2 and 3 did not follow the DiME account and/or the influencers on the list, thereby limiting the intended contrast between arms. Fidelity to arm 3 was especially low. It is possible that the assignment (to follow the campaign account and their choice of influencers) was overly complicated or time-consuming, leading participants to choose one or the other. Although two as-treated analyses confirmed findings from primary analyses, concordance between the two measures was low. Self-reported fidelity may overestimate exposure due to social desirability, whereas follow data may underestimate exposure due to changing handles or a tendency to view content without following.

Exposure intensity also varied among participants and represents a second aspect that is difficult to control on social media because it is influenced by the quantity of competing content and nuanced patterns of social media use. Most participants reported that they did not actively search for DiME content; instead, they relied on videos appearing organically in their feeds, increasing the possibility that content was missed. Several participants also reported following DiME accounts for only part of the study period. Regarding arm 3, mega-influencers posted only once, whereas micro-influencers reposted at variable rates; it is possible that their posts or shares were missed altogether. It is unknown what dose is required to produce changes in perceived stigma, and it is possible that, for less intensive exposure, the optimal duration exceeds the three-month frequency of this campaign. Finally, although this did not emerge from the qualitative data, participants may have been reluctant to follow an account that disseminated HIV-related content, particularly early in the campaign, when the overall number of followers was small. All in all, because low fidelity and variable exposure may have introduced misclassification and diluted differences between study arms, we are unable to determine whether the lack of observed change reflects an absence of effect or a combination of study design limitations and external factors.

The limitations of our study design, including variable exposure to content, reflect those of social media as a medium for message delivery. These challenges underscore the complexities of assessing the effects of social media campaigns on perceptions and behavior and align with prior research highlighting the difficulty of attributing outcomes to campaigns, which, by their nature, cannot be implemented in isolation.^31,32^ Alternative study designs may allow for greater control over some of these factors. For example, in a separate study, we tested four specific pieces of content among young people in Lima using a randomized design and a simulated social media feed.^33^ While we found that three of the four pieces were associated with a short-term reduction in stigma relative to control content, that study could not assess the real-world impact of the content delivered as part of an ongoing social media campaign.

Interestingly, we observed a change in the Concerns with Public Attitudes subscale across all study arms. While this finding may be due to chance or an underlying temporal trend, it is also possible that the campaign reduced viewers’ concerns. The campaign purposefully employed Peruvian influencers and young Peruvian actors to create content that normalized discussions around HIV and debunked myths about living with HIV.

Despite the inherent difficulties of quantitatively assessing the impact of social media campaigns, our qualitative findings support the role of locally tailored campaigns in combating HIV-related stigma. Future research should explore strategies to maximize engagement and exposure, evaluate the intervention across diverse populations, and integrate measures of exposure and impact to better evaluate its effectiveness.

## Authors’ Contributions

Conceptualization: MF (lead), RE (equal), JG (equal), KK (equal), MW (supporting), AN (supporting)

Data curation: MC (lead), MW (equal), YS (supporting), SR (supporting), AN (supporting)

Formal analysis: SR (lead), YS (equal), AN (supporting)

Funding acquisition: MF (lead), RE (supporting), JG (supporting), KK (supporting), MW (supporting) Investigation: all authors

Methodology: MF (lead), RE (equal), JG (equal), KK (equal), MW (equal)

Project administration: MW (lead)

Supervision: MF (lead), RE (equal)

Writing – Original draft preparation: SR (lead), MF (equal), AN (equal)

Writing – Review and editing: all authors

All authors have read and approved the final manuscript.

## Acknowledgements

The authors are indebted to the Socios En Salud Youth Advisory Board for their careful review of all aspects of the study. We would also like to thank Jelina Chavez and Rosario Peña for their work collecting quantitative data.

## Data Availability

The datasets generated or analyzed during this study are available from the corresponding author on reasonable request.

## Supporting Information

Supporting Information File 1: Qualitative interview guide and codebook

Supporting Information File 2: COREQ checklist

Supporting Information File 3: Supplemental Tables

