## Supplementary Material for "Impact of a Social Media Campaign on HIV-Related Stigma among Young Adults Living with HIV in Lima, Peru: A Sequential Explanatory Mixed Methods Study"

**<u>Appendix 1</u>**

**Table S1: Post-Intervention Interview Guide and Codebook**

| Question | Code |
|---|---|
| Tell us a little about your use of social media in relation to health. | Use of social media for health |
| [If yes] Have you ever searched for or followed accounts related to HIV issues? | Use of social media for HIV information |
| Could you tell us what motivated you to look for information on social media? | Motivation for seeking HIV information on social media |
| What did you think when we invited you to participate in this project and asked you to follow our social media account? | Reaction to invitation |
| What affected your decision to follow the account, or to not follow it? | Reasons for following account<br>Reasons for not following account |
| **Followers/Watchers** | |
| What content do you remember having the most impact on you? Did it impact you for positive reasons or negative reasons? What was the content like? Why did it impact you? | Content: positive impact<br>Content: negative impact |
| Our campaign, DiME, posted different types of content, testimonials from young people living with HIV, informational videos with actors, and interviews on the street. Can you tell us about the type of content that most caught your attention and why? | Most impactful content |
| What other types of content would you have liked to see? | Recommendations: content |
| What do you think about the frequency with which we have been posting our content? | Content frequency |
| How would you have liked this frequency to be? | Recommendations: frequency |

| | |
|---|---|
| How willing were you to share some of the campaign videos? | Willingness to share content |
| What motivated you to share or not to share? | Motivations to share<br>Motivations not to share |
| **Influencers** | |
| At the beginning there was a list of influencers that we asked you to follow. Did you follow any of them? | Follow influencers |
| What factors made you decide to follow them? What factors made you decide not to follow them? | Motivations for following influencers<br>Motivations for not following influencers |
| From that list, some influencers shared our content and others created educational content about HIV. Did you ever see that content? | Campaign exposure influencers |
| If so, what did you think of it? | Impressions of influencer content |
| What do you think about the participation of influencers in the DIME campaign? | Opinion re: influencer involvement in DiME |
| **Impact (on groups with positive, negative, or no change)** | |
| What is your opinion upon knowing/hearing that you are in this group because you have shown (or not shown) changes in your level of stigma? Do you agree with this change (or lack of change) according to the questionnaire between the months of June and November? | Reaction to stigma change grouping |
| What life experiences might you have had between June and November that might have had an impact on the change (or lack thereof) in your level of stigma? | External influences on stigma |
| How much do you think your contact with the DIME campaign has had an influence? | Campaign impact: stigma |
| At the beginning and at the end of this project, you answered some questions in a survey about stigma. What did you think of these questions? | Impressions of evaluation tools |

| | |
|---|---|
| How did you feel when you filled out the questionnaire? | Feelings evoked by evaluation |
| If you were analyzing this campaign, what other questions would you have asked in the survey to understand whether the campaign had an impact in reducing stigma? | Recommendations: evaluation |
| After seeing the content, how do you think the campaign can affect aspects apart from stigma? | Campaign impact: other |
| How did the videos on our account make you feel? | Emotions evoked by campaign videos |
| How did the comments that people posted on our videos or on the influencers' videos make you feel? | Emotions evoked by campaign comments |
| What will you take away from this campaign? Will you take away any new information from this campaign? | Campaign takeaways |
| How do you think this campaign may have influenced the general population? | Campaign impact: general population |
| After watching the videos on our account, whether from the account itself or from the influencers, how do you feel they have influenced or not influenced the way you see HIV? Or perhaps the way you perceive this health condition? | Campaign impact: perception of HIV |
| What topics would you have liked to see included that you feel were not covered? | Recommendations: topics |
| Are there other aspects of the campaign that you would have changed to improve it? | Recommendations: general |
| **For those who did not follow** | |
| What were the main reasons why you did not follow our account? | Reasons for not following |

| | |
|---|---|
| What could we have done differently to motivate you to follow our account? | Recommendations: following |
| Even though you didn't follow us, did you ever see the videos on our account or the influencer videos? | Campaign exposure |
| What would you have liked our social networks to have had to capture your attention and encourage you to follow us? | Recommendations: general |

**Appendix 2**

**Table S2: Consolidated criteria for reporting qualitative studies (COREQ) checklist**

| **No. Item** | **Guide questions/description** | **Reported on Page #** |
|---|---|---|
| **Domain 1: Research team and reflexivity** | | |
| *Personal Characteristics* | | |
| 1. Inter viewer/facilitator | Which author/s conducted the interview or focus group? | 5 |
| 2. Credentials | What were the researcher's credentials? E.g. PhD, MD | 5 |
| 3. Occupation | What was their occupation at the time of the study? | 5 |
| 4. Gender | Was the researcher male or female? | 5 |
| 5. Experience and training | What experience or training did the researcher have? | 5 |
| *Relationship with participants* | | |
| 6. Relationship established | Was a relationship established prior to study commencement? | 5 |
| 7. Participant knowledge of the interviewer | What did the participants know about the researcher? e.g. personal goals, reasons for doing the research | N/A. Participants were not informed so as not to introduce social desirability bias into responses. |
| 8. Interviewer characteristics | What characteristics were reported about the interviewer/facilitator? e.g. Bias, assumptions, reasons and interests in the research topic | N/A |
| **Domain 2: study design** | | |
| *Theoretical framework* | | |
| 9. Methodological orientation and Theory | What methodological orientation was stated to underpin the study? e.g. grounded theory, discourse analysis, ethnography, phenomenology, content analysis | 6 |
| *Participant selection* | | |
| 10. Sampling | How were participants selected? e.g. purposive, convenience, consecutive, snowball | 5 |
| 11. Method of approach | How were participants approached? e.g. face-to-face, telephone, mail, email | N/A. Participants had already consented to participate in the quantitative research |

| | | |
|---|---|---|
| | | and had routine communication with study staff for this reason. |
| 12. Sample size | How many participants were in the study? | 5 |
| 13. Non-participation | How many people refused to participate or dropped out? Reasons? | N/A. Only those indicating willingness to participate in qualitative research were contacted. |
| *Setting* | | |
| 14. Setting of data collection | Where was the data collected? e.g. home, clinic, workplace | 5 |
| 15. Presence of non-participants | Was anyone else present besides the participants and researchers? | 5 |
| 16. Description of sample | What are the important characteristics of the sample? e.g. demographic data, date | N/A. Demographic data was included for the overall sample. |
| *Data collection* | | |
| 17. Interview guide | Were questions, prompts, guides provided by the authors? Was it pilot tested? | 5 |
| 18. Repeat interviews | Were repeat interviews carried out? If yes, how many? | 5 |
| 19. Audio/visual recording | Did the research use audio or visual recording to collect the data? | 5 |
| 20. Field notes | Were field notes made during and/or after the interview or focus group? | 5 |
| 21. Duration | What was the duration of the interviews or focus group? | 5 |
| 22. Data saturation | Was data saturation discussed? | 5 |
| 23. Transcripts returned | Were transcripts returned to participants for comment and/or correction? | 6 |
| **Domain 3: analysis and findings** | | |
| *Data analysis* | | |
| 24. Number of data coders | How many data coders coded the data? | 6 |
| 25. Description of the coding tree | Did authors provide a description of the coding tree? | 6, Table S1 in Multimedia Appendix 1 |
| 26. Derivation of themes | Were themes identified in advance or derived from the data? | 6 |

| | | |
|---|---|---|
| 27. Software | What software, if applicable, was used to manage the data? | 6 |
| 28. Participant checking | Did participants provide feedback on the findings? | 6 |
| *Reporting* | | |
| 29. Quotations presented | Were participant quotations presented to illustrate the themes/findings? Was each quotation identified? e.g. participant number | 12-15 |
| 30. Data and findings consistent | Was there consistency between the data presented and the findings? | 12-15 |
| 31. Clarity of major themes | Were major themes clearly presented in the findings? | 12-15 |
| 32. Clarity of minor themes | Is there a description of diverse cases or discussion of minor themes? | 12-15 |

### Appendix 3

**Table S1. Intention-to-treat difference in differences, comparing mean change in intervention arms to that in the control arm, adjusting for baseline imbalances across groups***

| Measure | Adjusted difference in difference, Arm 2 relative to control (95% CI) | p-value | Adjusted difference in difference, Arm 3 relative to control (95% CI) | p-value |
|---|---|---|---|---|
| **HIV Stigma Scale** | | | | |
| Disclosure concerns | -0.45 (-1.76, 0.85) | 0.49 | -0.53 (-1.86, 0.79) | 0.43 |
| Negative self-image | 0.07 (-1.53, 1.67) | 0.93 | 0.20 (-1.44, 1.83) | 0.81 |
| Concern with public attitudes | -0.57 (-1.91, 0.77) | 0.40 | 0.10 (-1.28, 1.48) | 0.89 |
| Overall stigma score | -0.95 (-3.98, 2.07) | 0.53 | -0.41 (-3.51, 2.70) | 0.80 |
| **Stigma Stress Scale** | | | | |
| Overall stigma stress | 0.34 (-0.62, 1.31) | 0.48 | -0.11 (-1.10, 0.87) | 0.82 |

*Adjusted for age, sex assigned at birth, sexual orientation, gender identity, education level, and months since diagnosis.

**Table S2. Baseline characteristics of study participants, stratified by campaign exposure, as determined by study team assessment of following (as-treated analysis)**

| **Characteristic** | **Control (N = 100) n (%)*** | **Campaign-exposed (N = 41) n (%)*** |
|---|---|---|
| **Age**, mean (SD) | 25.7 (2.8) | 25.8 (3.0) |
| **Sex assigned at birth** | | |
| Female | 25 (25.0) | 6 (14.6) |
| Male | 75 (75.0) | 34 (82.9) |
| Intersex | 0 (0.0) | 0 (0.0) |
| Other | 0 (0.0) | 1 (2.4) |
| **Peruvian** | 92 (92.0) | 31 (75.6) |
| **Highest level of education** | | |
| Primary | 4 (4.0) | 1 (2.4) |
| High school | 53 (53.0) | 22 (53.7) |
| Trade school | 25 (25.0) | 6 (14.6) |
| University | 18 (18.0) | 11 (26.8) |
| Master's degree or higher | 0 (0.0) | 1 (2.4) |
| **Sexual orientation** | | |
| Heterosexual | 37 (37.0) | 9 (22.0) |
| Homosexual | 52 (52.0) | 24 (58.5) |
| Bisexual | 9 (9.0) | 8 (19.5) |
| Pansexual | 1 (1.0) | 0 (0.0) |
| Other | 1 (1.0) | 0 (0.0) |
| **Gender identity** | | |
| Woman | 25 (25.0) | 7 (17.1) |
| Man | 67 (67.0) | 32 (78.0) |
| Transgender woman | 2 (2.0) | 2 (4.9) |
| Transgender man | 1 (1.0) | 0 (0.0) |
| Gender fluid | 1 (1.0) | 0 (0.0) |
| Non-binary | 3 (3.0) | 0 (0.0) |
| Other | 1 (1.0) | 0 (0.0) |
| **Months since HIV diagnosis**, median (Q1, Q3, min, max) (N = 140) | 67.6 (28.8, 87.6, 0.6, | 74.0 (43.6, 96.2, |
| **Taking antiretroviral treatment** | 97 (97.0) | 39 (95.1) |
| **Disclosed HIV diagnosis to $\geq$ 1 person** | 90 (90.0) | 37 (90.2) |
| **Experienced stigma or discrimination in the last 12 months** | 79 (79.0) | 33 (80.5) |
| **Self-description of health** | | |
| Excellent | 14 (14.0) | 11 (26.8) |
| Very good | 27 (27.0) | 12 (29.3) |
| Good | 38 (38.0) | 12 (29.3) |
| Regular | 20 (20.0) | 6 (14.6) |
| Bad | 1 (1.0) | 0 (0.0) |
| **Has a smartphone** | 96 (96.0) | 35 (85.4) |
| **Uses TikTok daily** (N = 134) | 76 (76.0) | 26 (63.4) |
| **Hours spent per day on TikTok**, median (Q1, Q3) (N = 140) | 4 (2, 5.2) | 3 (2, 5) |
| **Uses Instagram daily** (N = 138) | 53 (53.0) | 27 (65.9) |
| **Hours spent per day on Instagram**, median (Q1, Q3) (N = 138) | 2 (1, 3) | 2 (1, 3) |

*Unless otherwise noted.

**Table S3. Baseline characteristics of study participants, stratified by campaign exposure, as determined by participant self-report (as-treated analysis)**

| **Characteristic** | **Control (N = 31) n (%)*** | **Campaign-exposed (N = 110) n (%)*** |
|---|---|---|
| **Age**, mean (SD) | 25.2 (3.2) | 25.9 (2.7) |
| **Sex assigned at birth** | | |
| Female | 6 (19.4) | 25 (22.7) |
| Male | 25 (80.6) | 84 (76.4) |
| Intersex | 0 (0.0) | 0 (0.0) |
| Other | 0 (0.0) | 1 (0.9) |
| **Peruvian** | 29 (93.5) | 94 (85.5) |
| **Highest degree or level of education** | | |
| Primary | 2 (6.5) | 3 (2.7) |
| High school | 19 (61.3) | 56 (50.9) |
| Trade school | 5 (16.1) | 26 (23.6) |
| University | 5 (16.1) | 24 (21.8) |
| Master's degree or higher | 0 (0.0) | 1 (0.9) |
| **Sexual orientation** | | |
| Heterosexual | 8 (25.8) | 38 (34.5) |
| Homosexual | 19 (61.3) | 57 (51.8) |
| Bisexual | 2 (6.5) | 15 (13.6) |
| Pansexual | 1 (3.2) | 0 (0.0) |
| Other | 1 (3.2) | 0 (0.0) |
| **Gender identity** | | |
| Woman | 6 (19.4) | 26 (23.6) |
| Man | 24 (77.4) | 75 (68.2) |
| Transgender woman | 0 (0.0) | 4 (3.6) |
| Transgender man | 0 (0.0) | 1 (0.9) |
| Gender fluid | 1 (3.2) | 0 (0.0) |
| Non-binary | 0 (0.0) | 3 (2.7) |
| Other | 0 (0.0) | 1 (0.9) |
| **Months since HIV diagnosis**, median (Q1, Q3, min, max) (N = 140) | 60.1 (23.0, 80.7, 7.1, 288.1) | 72.2 (39.8, 98.4, 0.5, 282.8) |
| **Taking antiretroviral treatment** | 30 (96.8) | 106 (96.4) |
| **Disclosed HIV diagnosis to $\geq$ 1 person** | 30 (96.8) | 97 (88.2) |
| **Experienced stigma or discrimination in the last 12 months** | 25 (80.6) | 87 (79.1) |
| **Self-description of health** | | |
| Excellent | 5 (16.1) | 20 (18.2) |
| Very good | 10 (32.3) | 29 (26.4) |
| Good | 9 (29.0) | 41 (37.3) |
| Regular | 7 (22.6) | 19 (17.3) |
| Bad | 0 (0.0) | 1 (0.9) |
| **Has a smartphone** | 31 (100.0) | 100 (90.9) |
| **Uses TikTok daily** (N = 134) | 24 (77.4) | 78 (70.9) |
| **Hours spent per day on TikTok**, median (Q1, Q3) (N=140) | 3 (2, 5) | 3 (2, 5.5) |
| **Uses Instagram daily** (N = 138) | 16 (51.6) | 64 (58.2) |

| | | |
|---|---|---|
| **Hours spent per day on Instagram**, median (Q1, Q3) (N=138) | 2 (1, 3) | 2 (1, 3) |

*Unless otherwise noted.

**Table S4. Mean HIV Stigma and Stigma Stress scores and change scores (as-treated analysis; campaign exposure determined by study team assessment)**

| Measure | Control (N= 31) | | | Campaign-exposed (N = 110) | | | |
|---|---|---|---|---|---|---|---|
| | Pre mean (sd) | Post mean (sd) | Mean change (95% CI) | Pre mean (sd) | Post mean (sd) | Mean change (95% CI) | p-value* |
| **HIV Stigma Scale** | | | | | | | |
| Disclosure concerns | 15.4 (3.8) | 15.1 (3.6) | -0.27 (-1.02, 0.48) | 15.3 (3.9) | 14.5 (3.8) | -0.80 (-1.98, 0.37) | 0.45 |
| Negative self-image | 11.1 (4.3) | 11.6 (4.5) | 0.47 (-0.45, 1.39) | 11.1 (4.7) | 11.0 (4.5) | -0.07 (-1.33, 1.18) | 0.51 |
| Concern with public attitudes | 14.4 (4.5) | 13.8 (3.7) | -0.67 (-1.44, 0.10) | 14.9 (3.1) | 13.7 (3.5) | -1.20 (-2.46, 0.07) | 0.47 |
| Overall stigma score | 40.9 (10) | 40.4 (9.1) | -0.47 (-2.12, 1.18) | 41.3 (8.0) | 39.2 (9.2) | -2.07 (-4.76, 0.61) | 0.30 |
| **Stigma Stress Scale** | | | | | | | |
| Overall stigma stress score | -2.4 (2.7) | -2.2 (2.3) | 0.27 (-0.32, 0.85) | -2.5 (2.4) | -2.7 (2.9) | -0.16 (-1.09, 0.76) | 0.43 |

*Two-sample paired t-test.

**Table S5. As-treated difference in differences, comparing mean change in campaign-exposed individuals to that in the control arm, adjusting for baseline imbalances across groups* (campaign exposure determined by study team assessment)**

| Measure | Adjusted difference in difference, campaign-exposed, relative to control (95% CI) | p-value |
|---|---|---|
| **HIV Stigma Scale** | | |
| Disclosure concerns | 0.16 (-1.15, 1.46) | 0.81 |
| Negatives self-image | 1.32 (-0.26, 2.91) | 0.10 |
| Concern with public attitudes | -0.57 (-1.91, 0.77) | 0.40 |
| Overall stigma score | 0.91 (-2.12, 3.93) | 0.56 |
| **Stigma Stress Scale** | | |
| Overall stigma stress score | -0.30 (-1.27, 0.66) | 0.53 |

*Adjusted for baseline covariate differences in age, sex assigned at birth, sexual orientation, gender identity, education level, and months since diagnosis.

**Table S6. Mean HIV Stigma and Stigma Stress scores and change scores (as-treated analysis; campaign exposure determined by self-report)**

| Measure | Control (N= 31) | | | Campaign-exposed (N = 110) | | | |
|---|---|---|---|---|---|---|---|
| | Pre mean (sd) | Post mean (sd) | Mean change (95% CI) | Pre mean (sd) | Post mean (sd) | Mean change (95% CI) | p-value* |
| **HIV Stigma Scale** | | | | | | | |
| Disclosure concerns | 16.1 (3.1) | 15.3 (2.9) | -0.87 (-1.88, 0.14) | 15.1 (4.0) | 14.8 (3.8) | -0.30 (-1.06, 0.46) | 0.46 |
| Negative self-image | 11.3 (4.1) | 10.8 (4.2) | -0.42 (-1.78, 0.95) | 11.0 (4.5) | 11.6 (4.6) | 0.52 (-0.36, 1.39) | 0.30 |
| Concern with public attitudes | 14.4 (3.8) | 14.1 (2.5) | -0.35 (-1.52, 0.81) | 14.6 (4.2) | 13.6 (3.9) | -0.95 (-1.73, 0.18) | 0.45 |
| Overall stigma score | 41.8 (8.7) | 40.2 (7.6) | -1.65 (-4.13, 0.84) | 40.8 (9.7) | 40.0 (9.5) | -0.74 (-2.40, 0.93) | 0.60 |
| **Stigma Stress Scale** | | | | | | | |
| Overall stigma stress score | -2.3 (2.4) | -1.9 (2.2) | 0.35 (-0.62, 1.33) | -2.5 (2.7) | -2.4 (2.5) | 0.08 (-0.49, 0.65) | 0.65 |

*Two-sample paired t-test.

**Table S7. As-treated difference in differences, comparing mean change in campaign-exposed individuals to that in the control arm, adjusting for baseline imbalances across groups* (campaign exposure determined by self-report)**

| Measure | Adjusted difference in difference, campaign-exposed, relative to control (95% CI) | p-value |
|---|---|---|
| **HIV Stigma Scale** | | |
| Disclosure concerns | -0.56 (-1.75, 0.62) | 0.35 |
| Negative self-image | -0.44 (-1.91, 1.02) | 0.55 |
| Concern with public attitudes | -0.31 (-1.54, 0.92) | 0.62 |
| Overall stigma score | -1.35 (-4.12, 1.42) | 0.34 |
| **Stigma Stress Scale** | | |
| Overall stigma stress score | -0.34 (-1.22, 0.54) | 0.44 |

*Adjusted for age, sex assigned at birth, sexual orientation, gender identity, education level, and months since diagnosis.

**Table S8. Mean change HIV Stigma and Stigma Stress scores, across all study arms (N = 141)**

| **Measure** | **Mean change (95% CI)** | **p-value** |
|---|---|---|
| **HIV Stigma Scale** | | |
| Disclosure concerns | -0.43 (-1.05, 0.20) | 0.18 |
| Negative self-image | 0.31 (-0.43, 1.05) | 0.41 |
| Concern with public attitudes | -0.82 (-1.47, -0.17) | 0.01* |
| Overall stigma score | -0.94 (-2.33, 0.46) | 0.19 |
| **Stigma Stress Scale** | | |
| Overall stigma stress score | 0.14 (-2.33, 0.46) | 0.57 |
